# Transmission of Respiratory Infectious Diseases between Neighboring Cities using Agent-based Model and Compartmental Model

**DOI:** 10.1101/2020.06.24.20138818

**Authors:** Christian Alvin H. Buhat, Destiny SM. Lutero, Yancee H. Olave, Monica C. Torres, Jomar F. Rabajante

**Affiliations:** UPLB Biomathematics Research Cluster, and UPLB Quantitative Management and Decision Science Research Cluster, Institute of Mathematical Sciences and Physics, University of the Philippines Los Baños, 4031 Laguna, Philippines; Faculty of Education, University of the Philippines Open University, 4031 Laguna, Philippines

**Keywords:** infectious disease, disease transmission, respiratory infections, agent-based model, compartmental model

## Abstract

We formulate an agent-based model and a compartmental model (SEIR) that simulate the spread of a respiratory infectious disease between two neighboring cities. We consider preventive measures such as implementation of social distancing and lockdown in a city, as well as the effect of protective gears or practices. The chance of travelling to another city and within the city during lockdown, and initial percentage of exposed and infected individuals on both cities influence the increase in the number of newly-infected individuals on both models. Our simulations show that (i) increase in exposed individuals results in increase in number of new infections, hence the need for increased testing-isolation efforts; (ii) protection level of 75-100% effectiveness impedes disease transmission; (iii) travelling within city or to other city can be an option given that strict preventive measures (e.g., non-pharmaceutical interventions) are observed; and (iv) the ideal set-up for neighboring cities is to implement lockdown when there is high risk of disease local transmission while individuals observe social distancing, maximizing protective measures, and isolating those that are exposed. The results of the agent-based and compartmental models show similar qualitative dynamics; the differences are due to different spatio-temporal heterogeneity and stochasticity. These models can aid decision makers in designing infectious disease-related policies to protect individuals while continuing population movement.

## 1 Introduction

The spread of respiratory infectious diseases has brought havoc worldwide. In the last century, the worst global outbreaks of influenza happened in 1918, 1957, 1968 and 2009, with the 1918 pandemic resulting in up to 50 million deaths [17]. Shortly after the turn of the millennium, Severe acute respiratory syndrome coronavirus (SARS-CoV) was identified to be caused by a novel coronavirus and has caused an epidemic in 29 countries and regions [18]. In early January 2020, another respiratory illness caused by the coronavirus called SARS-CoV-2 (more commonly known as Coronavirus Disease 2019 or COVID-19) was identified and the outbreak of this disease was later announced by the World Health Organization (WHO) as a global pandemic [5, 7, 22].

The worldwide spread of COVID-19 has driven change in mobility. According to studies, population movement is a major driver of transmission for infections like COVID-19 [8, 16, 20]. Such movements are necessary for aspects such as economy but is not possible during a pandemic until a vaccine against the disease is developed [14]. Before a vaccine becomes available worldwide, non-pharmaceutical interventions (NPI) are the mainstay to control the spread of COVID-19 [2, 6, 9, 12]. Different forms of community lockdowns were enforced in affected areas as means to delay the spread of infection [2, 12, 15]. NPIs during community lockdowns often include social distancing in public spaces, closure of schools and workplaces, limiting public transportation within and between communities, and reducing sizes of gatherings [2, 12]. These measures may need to be implemented for months to control the spread of infections like COVID-19 [13]. However, prolonging lockdowns may lead to unnecessary detriment to the economy [6, 15]. Thus, intermittent NPIs are thought to be better options [3, 6, 9]. One study suggests dynamic cycles of 50-day suppression followed by 30-day relaxation over an 18-month period or until a vaccine arrives [6] while in another, a cyclic schedule of 4-day work and 10-day lockdown coupled with certain conditions suppresses the epidemic without paralyzing the economy [19].

To study such interactions, several studies used mathematical models to simulate the spread of diseases. Oftentimes, two models are used to study the transmission of these diseases: differential equation-based models (EBM), and agent-based models (ABM) [23]. An agent-based model that factored in population, area, vaccination rates, and age structure from openly-available data was used to observe transmission of an airborne infectious disease [11]. Another used epidemic diffusion model to simulate transmission via road systems [21], while [4] used SIR epidemic model to observe transmission dynamics in two cities through transport.

In this study, we present our analyses of the roles of NPIs (implementation of social distancing and lockdown in a city/-community) and key parameters (protective gears and practices, travelling to a nearby city, going out during lockdown, and initial percentage of infectious individuals) on the transmission of respiratory infectious diseases between neighboring cities. We use an ABM and an SEIR model, with COVID-19 as an example of a respiratory infectious disease in our simulations. The *Computational Models* section gives information on how simulations work, buttons’ functions, and how simulations can be performed on each of the models to be used. The *Simulation Results* section discusses the impact of initial percentage of exposed and infected individuals, protection level, travel rate, and go-out rate during lockdown. The *Conclusion* section summarizes the key findings of the study. The *Limitations* section details the opportunities for further improvement of the study.

## 2. Computational Models

### 2.1. Agent-based Model

We first use an agent-based model (ABM) to simulate the transmission of a respiratory infectious disease between two neighboring cities using the NetLogo simulation environment. ABM is a micro-scale model used to demonstrate movements and interactions among agents in a complex system with the purpose of simulating their behavior [10]. It has been extensively used in various fields such as in biology to study the population dynamics and simulate the interaction among the individuals in the population [1].

The model simulates the spread of a respiratory infectious disease, such as (COVID-19) between two neighboring cities. The model explores the effects of factors such as protection of individuals against infection, chance of an individual travelling to another city, chance of an individual leaving their house during lockdown, initial number of exposed individuals, and initial number of infected individuals per city. We determine these effects under the presence or absence of social distancing protocol, and presence/absence of citywide lockdowns.

Individuals are initiated and placed randomly in each city. They may or may not travel to the neighboring city depending on their travel rate. In the presence of lockdown, an individual is expected to stay put but may leave their position depending on the going out rate. Their position will determine if they will be infected or not. A person within 3 meters of an infectious individual will have a 25% chance of being exposed, a person within 2 meters of an infectious individual will have a 50% chance of being exposed while a person within 1 meter of an infectious individual will have a 75% chance of being exposed. Exposed individuals may or not may develop into an infected individual depending on the protection. Protection (handwashing, use of alcohols, sanitizers) is the chance for an exposed individual to be an unexposed or uninfected individual again, while if the person fails to protect him or herself, the person will become infected. Infected individuals will then be infectious to those people surrounding them for 14 days.

The **SETUP** button creates individuals according to the parameter values chosen by the user and these individuals are divided equally among each cities. Each individual has a chance of initially being exposed depending on the INIT-EXP value and being infected depending on the INIT-INF value (INIT-INF1 for city 1, and INIT-INF2 for city 2). Once the model has been set up, the **GO** button starts the model simulations. GO starts the model and runs it continuously until: (1) all individuals are infected, or (2) no more infections can occur. Each time step can be considered as a day or any suitable time unit will do.

The following are the summary of the variables for the model:

| Variable | Description | Default Value |
| --- | --- | --- |
| INITIAL-HUMAN-POPULATION | Initial number of individuals to be divided equally in both cities | 100 |
| INFECTIOUS-DAYS | Number of days (ticks) for an infected individual to be infectious to other unexposed or uninfected ones | 14 days (ticks) |
| PROTECTION | Chance for an exposed individual to be back to being susceptible again | 0 to 100% |
| TRAVELRATE | Chance for an individual to travel to the neighboring city | 0 to 100% |
| GOOUT-RATE | Chance for an exposed individual to move from his/her position during lockdown | 0 to 100% |
| INIT-EXP | Initial percentage of exposed individuals in both cities | 0 to 75% |
| INIT-INF1 | Initial percentage of infected individuals in City 1 | 0 to 75% |
| INIT-INF2 | Initial percentage of infected individuals in City 2 | 0 to 75% |
| SOCIAL-DISTANCING | if this switch is on, all individuals will try to stay 1m apart | Yes/No |
| LOCKDOWN | If this switch is on, all individuals, will stay in his/her city and will only move based on the GOOUT-RATE | Yes/No |

### 2.2. SEIR Model

We consider an Extended Susceptible-Exposed-Infected-Recovered (SEIR) compartmental model to study the dynamics and transmission of respiratory infectious disease between two neighboring cities. Similar to the Agent-based model, the model simulates the spread of a respiratory infectious disease, and explores the similar factors under the same preventive measures used in the ABM.

In the model, the residents of City 1 and City 2 are compartmentalized to Susceptible, Exposed, Infected, and Recovered. We note that classes with subscript 1 are from City 1 and compartments with subscript 2 are from City 2. The susceptible residents are grouped into those that cannot move around the city (*S*_1_ and *S*_2_) and the residents that may go out (*S g*_1_ and *S g*_2_). The other classes include exposed individuals (*E*_1_ and *E*_2_), infected individuals (*I*_1_ and *I*_2_), and the removed individuals (*R*_1_ and *R*_2_) who are no longer infectious. Part of susceptible individuals (*S*) may leave their location and are transferred to *S*_*g*_ at rate *γ*. A portion *λ* of those that can move around will be immobile. We let *β* as the exposure rate of the individuals to the respiratory infectious disease. The exposure rate is the number of new exposed individuals caused by an infectious individual per unit time. The rate at which an exposed individual in either of the cities become vulnerable or susceptible to the transmission of the virus is given by 1 − *µ*. Exposed individuals become infected with the disease at a rate of *α* for both City 1 and City 2 residents. Individuals in each city travel to another city at rate *τ*. Moreover, infected individuals become non-infectious and are transferred to the removed class *R* at rate *δ*. The table below shows the values that were used in the simulations performed in this study.

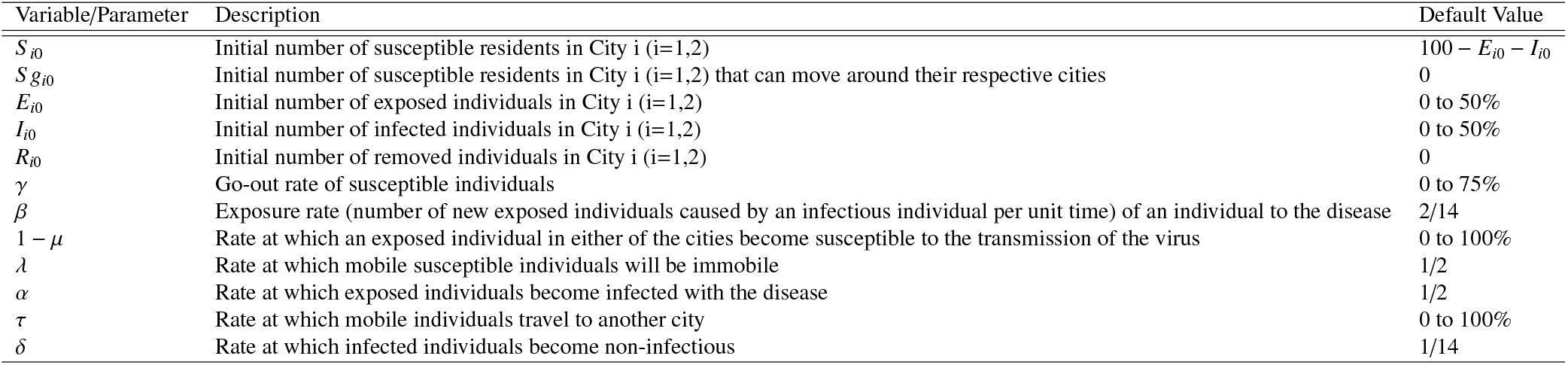

The dynamics in the extended SEIR Compartment Model is described by the following equations.

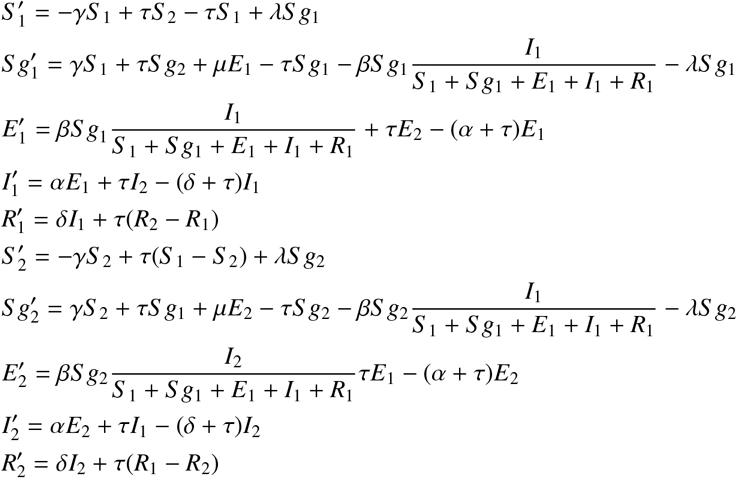

## 3. Results and Discussion

We vary key parameters in both the agent-based model (ABM) and the extended SEIR model. We determine the effect of each parameter on the number of newly infected individuals as the number of initial infected population on both cities increase. We observe their behaviour under different scenarios (with or without social distancing measure, and with or without lockdown protocol) for up to 500 ticks.

### 3.1. Initial Number of Exposed Individuals

With a fixed percentage of go out rate, protection, and travel rate, we vary the initial number of exposed individuals and initial infected population of both cities. We simulate scenarios where both lock down and social distancing are present, one of them is off, and both are not applied.

It is shown in the ABM heatmaps from Figure 3 that with or without lockdown and social distancing, a larger infected population is observed if the initial exposed population is increased while keeping the other parameters constant. This implies that lockdown and social distancing will be useless if there is still a significant exposed population. Testing will play an important role here since identifying the infected ones and isolating them will lessen the exposed individuals. Comparing the scenarios when the initial exposed is 25% of the population, the case with no lockdown and with social distancing has a slightly bigger infected population as compared to the case with lockdown and no social distancing. This implies that a minimum travel of individuals is still better than social distancing. It can also be noticed in each scenario that a larger number of initial infected in both cities (high prevalence) will result in a further greater infected population. Similar to the ABM simulations, heatmaps from the SEIR simulations show that in each scenario, a greater number of initial infected in both cities will result in a larger infected population. Furthermore, as initial exposed population increases, infected population increases as well. This implies that lockdown and social distancing may not be effective in controlling the spread of infection if there is still a significant exposed population. A lesser infected population is observed if social distancing is enforced as compared when lockdown is implemented. This means that moving within the city will not worsen the situation as long as strict physical distancing is followed.

**Figure 1:**
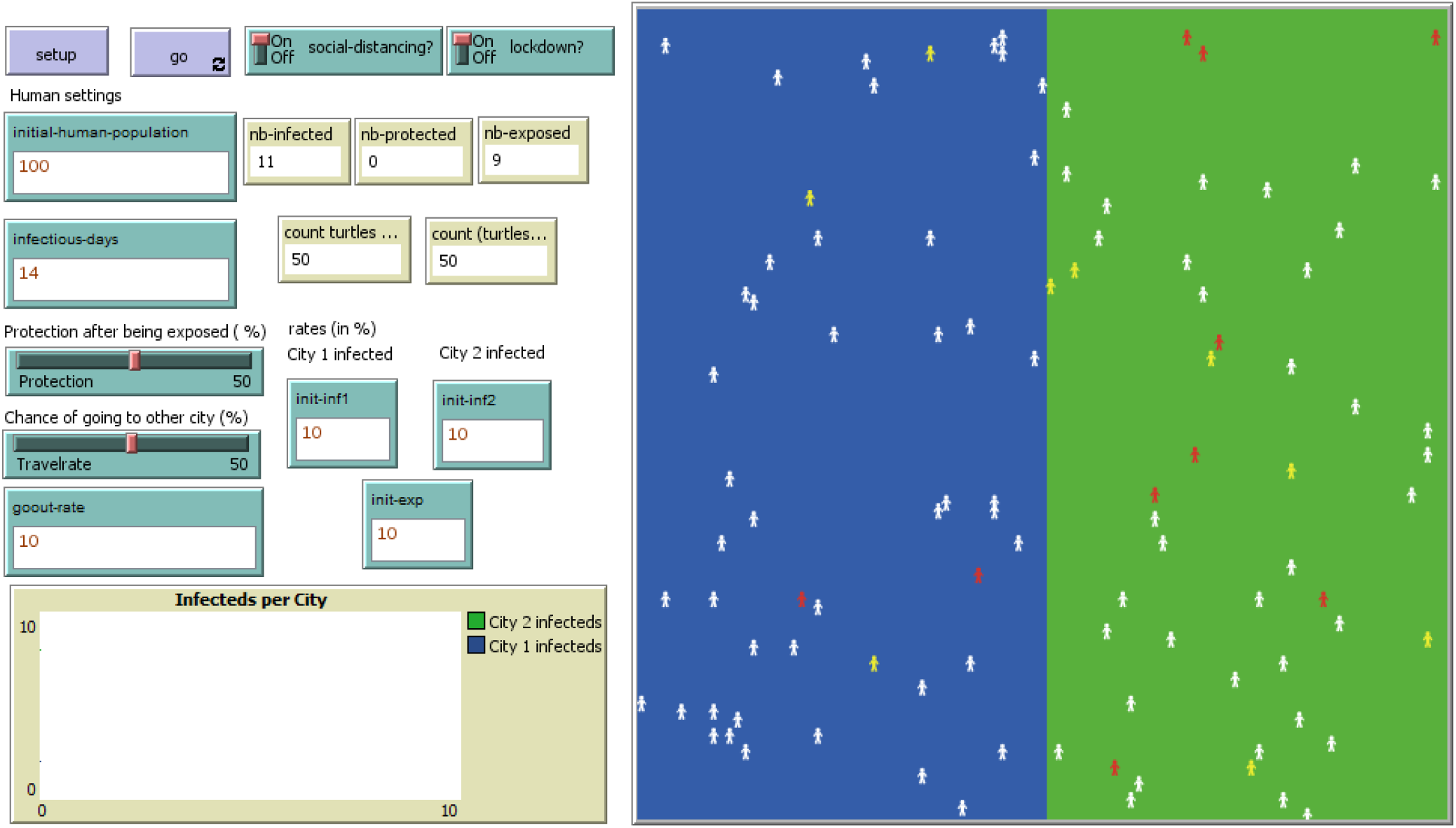
NetLogo simulation environment of transmission of respiratory infectious disease between two neighboring cities

**Figure 2:**
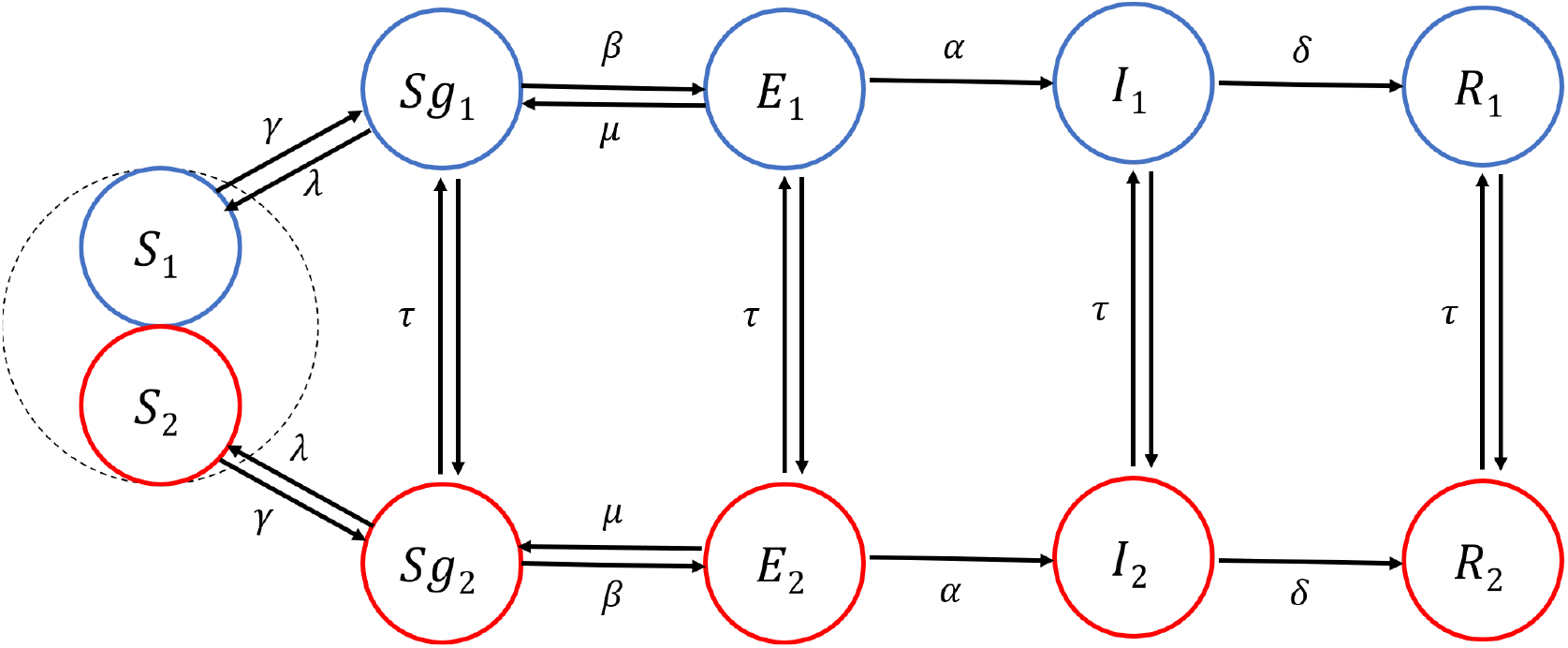
Extended Susceptible-Exposed-Infected-Recovered Model framework of transmission of respiratory infectious disease between two neighboring cities

**Figure 3:**
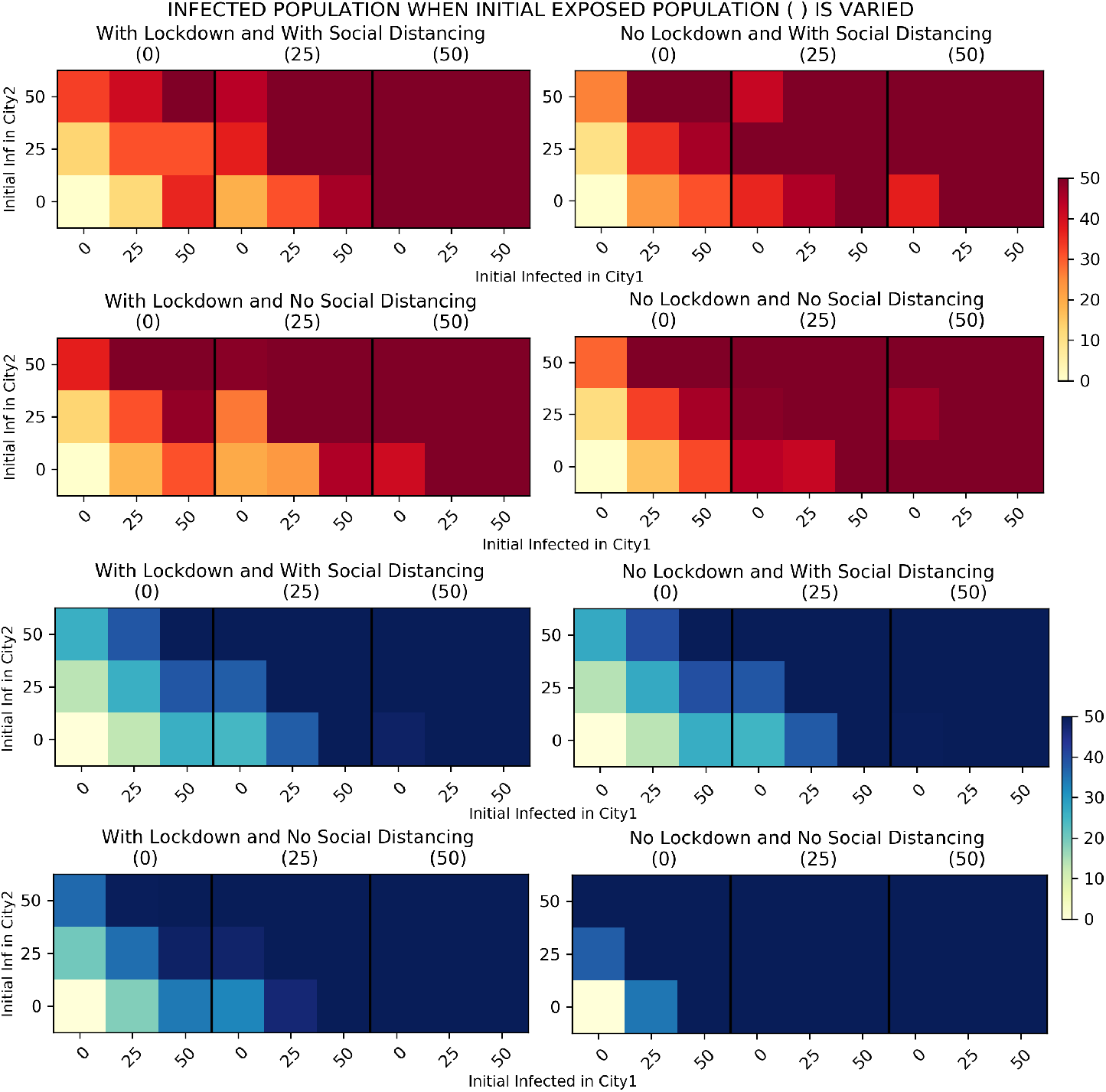
Resulting final number of infected individuals, from ABM simulations (red colorway) and SEIR model simulations (blue colorway), given various initial percentage of exposed individuals with different initial percentage of infected individuals in City 1 (in %) and the initial percentage of infected individuals in City 2 (in %), with protection rate = 25%, travel rate = 25%, and goout-rate = 25% (during lockdown)

An obvious difference in the result of the two models is that social distancing has a better effect in the extended SEIR model compared to lockdown (see blue heatmaps upper right and lower left subfigures). This is in contrary with the result in ABM. This inconsistency is anticipated since interactions between individuals is removed in the EBM. Furthermore, disease progression in the SEIR is similar for all individuals. This property resulted in an almost perfect employment of social distancing in the SEIR model when it is ‘on’, i.e., only small part of the population will transfer to the Exposed compartment. On the other hand, applying social distancing in the ABM involves stochasticity, that is, social distancing will only lessen the probability of and will not totally eliminate exposure of each agent to the infection. We also note that this probability varies for each agent.

### 3.2. Protection against Infection

In the previous subsection, we saw that a larger initial exposed population increases the number of infections. Now we look at the effect of varying the protection rate from 0 to 100 percent, with a fixed percentage of initial exposed individuals, travel rate and go out rate.

Simulation results from ABM and SEIR model from Figure 4 show that having individual protection is necessary to lessen the number of infections in two cities. Based on the results of ABM, additional NPIs are needed when protection are not 100% effective. It can also be seen that imposing a lockdown coupled with protection is better than social distancing with protection especially when the protection level is very low. These are not the case when the SEIR model is used. It is depicted from the heatmaps of the SEIR model simulations that increasing the protection fully is not enough to reduce the number of infections. When additional interventions are introduced, it is shown that protection with social distancing is better than protection with lockdown. The results of having social distancing with lockdown is almost the same when there is no lockdown. This is again due to the employment of almost perfect rate of physical distancing when it is ‘on’ and the homogeneous mixing of individuals. We can also observe from the SEIR heatmaps that even though these control measures are observed, the number of infections is still high when there are at least 25% infected individuals present in each cities. This implies that early detection of infected individuals and isolating them together with contact tracing are very important to keep the number of infected individuals at a minimum.

**Figure 4:**
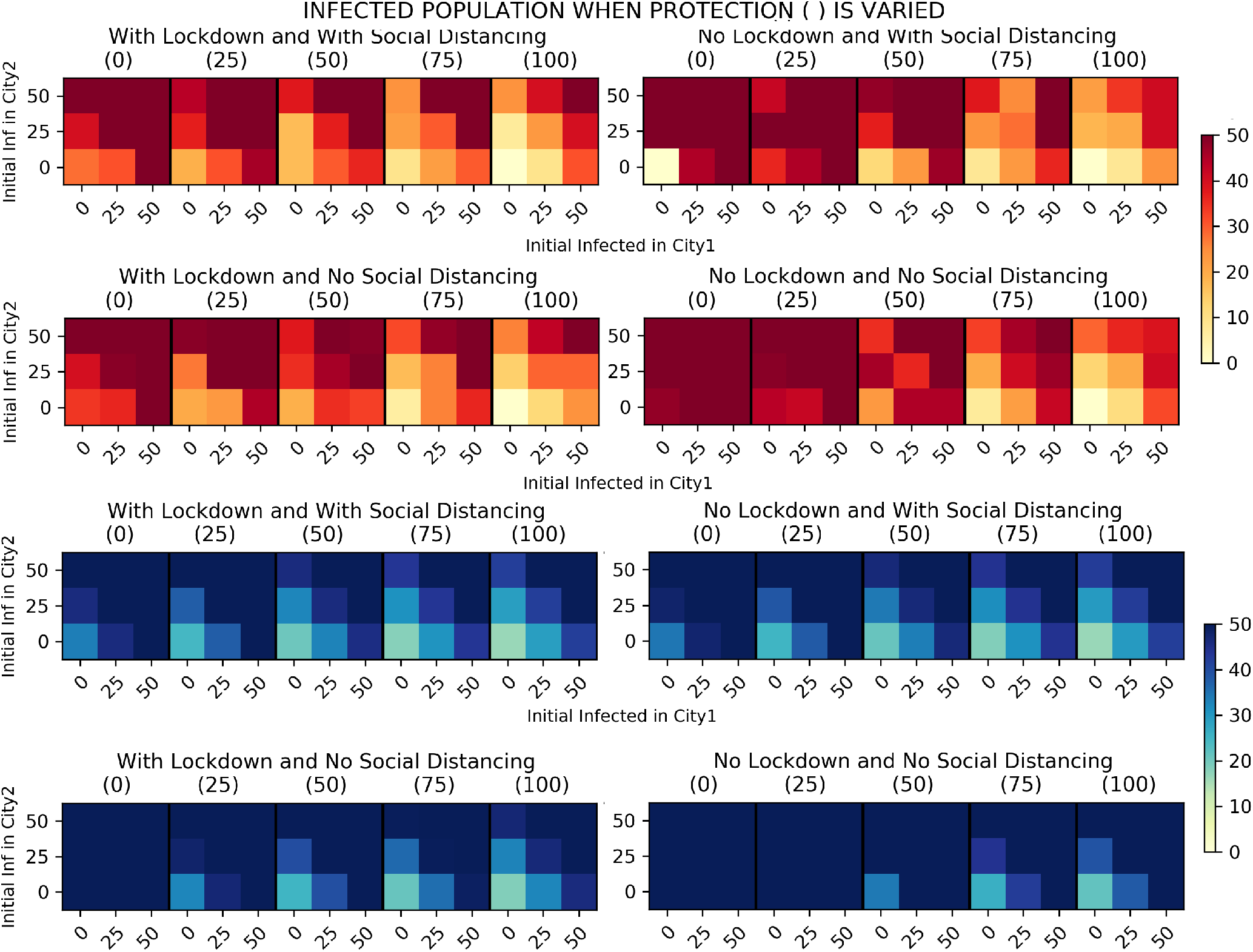
Initial size of the exposed population is varied. Resulting final number of infected individuals, from ABM simulations (red colorway) and SEIR model simulations (blue colorway), given various protection rate of individuals with different initial percentage of infected individuals in City 1 (in %) and the initial percentage of infected individuals in City 2 (in %), with initial exposed percentage = 25%, travel rate = 25%, and go outrate = 25% (during lockdown)

#### 3.3. Effect of Travelling between Cities

In the previous subsection, we observed that as protection increases, the number of infected individuals between two cities decreases. Now, we look at the effect of varying the travel rate from 0 to 100 percent with a fixed percentage of initial exposed individuals, protection rate and go out rate.

Both simulations from ABM and SEIR model show that increasing the travel rate has no significant effect in all scenarios given that the other parameters are all constant, as shown in Figure 5. Individuals can travel between two cities if there is at most 25% infected individuals present in both cities, protection against infection is present, and at least social distancing is observed. Imposing a lockdown, social distancing or both is necessary to minimize the number of infections as we can observe from the heat maps. In addition, SEIR heatmaps show that infected population is greatest when social distancing is not employed. In all four scenarios, changes in travel rate has no effect to the infected population.

**Figure 5:**
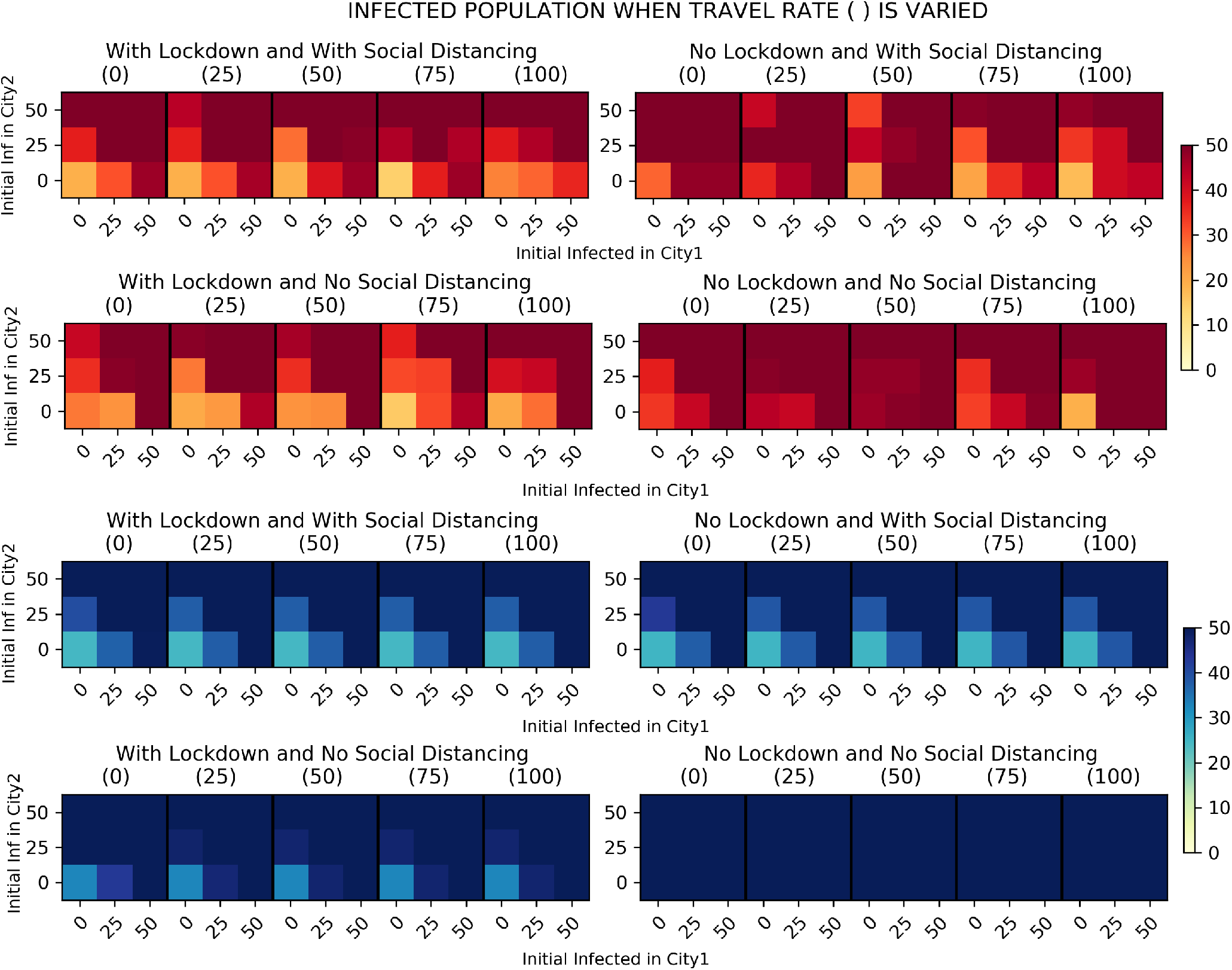
Protection level is varied. Resulting final number of infected individuals, from ABM simulations (red colorway) and SEIR model simulations (blue colorway), given various travel rate of individuals with different initial percentage of infected individuals in City 1 (in %) and the initial percentage of infected individuals in City 2 (in %), with initial exposed percentage = 25%, protection rate = 25%, and goout-rate = 25% (during lockdown)

### 3.4. Effect of Travelling within the city during Lockdown

With a fixed percentage of travel rate, protection, and initial exposed individuals, we vary the go out rate and initial infected population of both cities. We test scenarios such as lockdown within the city and application of social distancing.

In both models shown in Figure 6, infected population is greater when social distancing is not practiced. Go out rate has little (ABM) to no effect (SEIR) in terms of infected population when social distancing is in effect. This means that if the population follows proper social distancing protocols, number of infected can be kept at a minimum. On the other hand, the effect of go out rate is evident when there is no social distancing. Thus, in situations or places where social distancing is difficult to implement, number of mobile individuals should be reduced.

**Figure 6:**
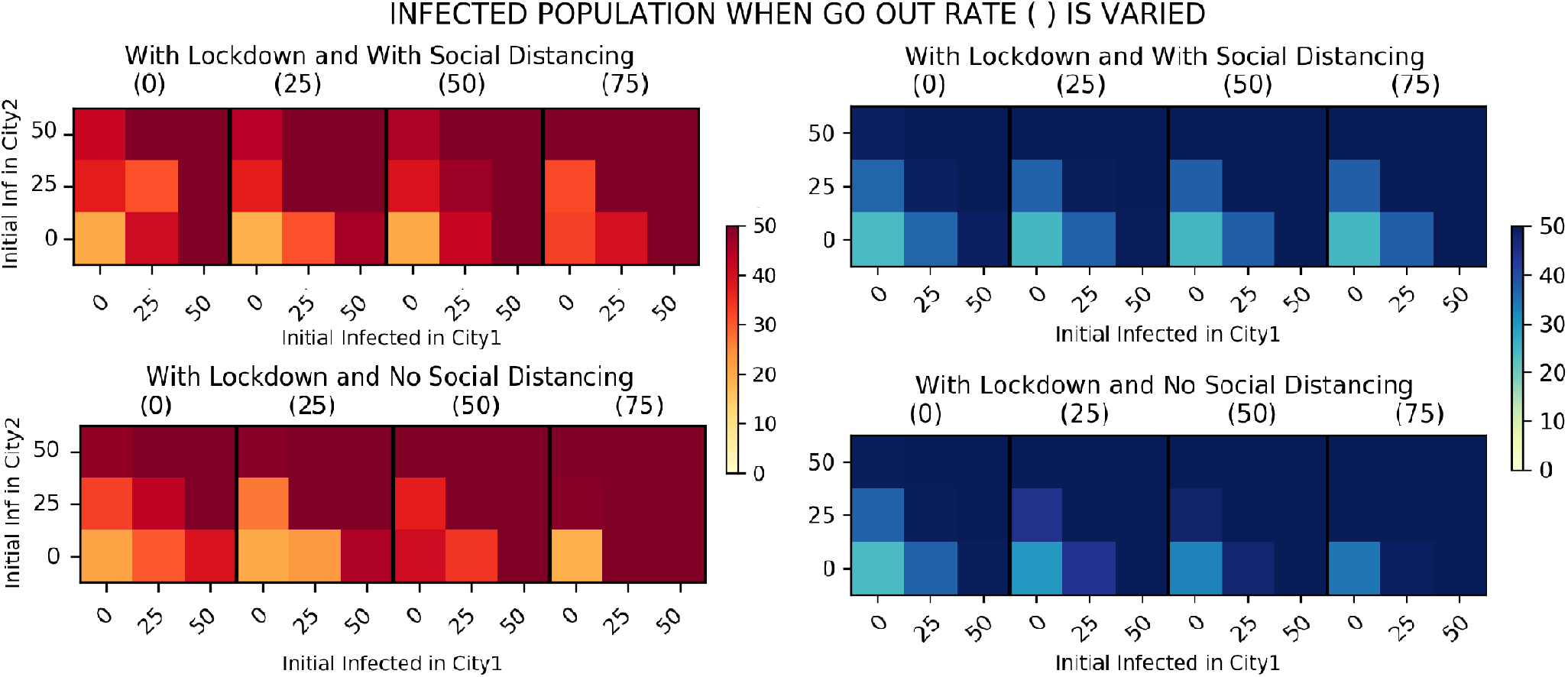
Go out rate is varied. Resulting final number of infected individuals, from ABM simulations (red colorway) and SEIR model simulations (blue colorway), given various go out rate of individuals during lockdown with different initial percentage of infected individuals in City 1 (in %) and the initial percentage of infected individuals in City 2 (in %), with initial exposed percentage = 25%, protection rate = 25%, and travel rate = 25%

Comparing the upper red and the upper blue heatmaps, go out rate has an effect in the ABM and has very little to no effect in the extended SEIR model. This disagreement is again a consequence of the stochasticity (applied in ABM) in the exposure rate when social distancing is ‘on’. Agents in the ABM will more likely to interact as go out rate is increased, which may heighten the infected population. On the other hand, since individuals in each group in EBM have the same behavior or follow the same pattern, go out rate had no impact when physical distancing is applied. In other words, since a high level of social distancing is set in our simulations, almost all individuals who move around will perfectly follow physical distancing.

## 4. Conclusion

In this study, an agent-based model and an extended SEIR model were used to determine the effects of non-pharmaceutical interventions and other factors in the spread of a respiratory infection between two cities. We considered all combinations of scenarios involving the implementation of social distancing and lockdown among individuals. We varied values for key parameters, such as protection level of individuals, travel rate, go out rate, and initial percentage of exposed and infected individuals, affecting the transmission of respiratory infectious disease between two cities. We then analyzed the effect of these parameters to the number of individuals that have been newly infected from the transmission of the disease.

Simulations for both ABM and SEIR models show that isolation of infected individuals is necessary to reduce the exposed individuals since greater exposure means greater chances of infection. Mass testing will play an important role here since isolating the infected ones will lessen the exposed individuals. Protection measures for individuals against infection is another vital step in reducing the chance of infection. Protection level of at least 75% together with practice of social distancing and lockdown is recommended (and maximized to 100% if possible) to lessen disease transmission between cities. Frequent washing or cleaning of hands, wearing of mask, and other protective measures should then be a habit for each individual. During lockdown, going out rate matters only when social distancing is not practiced. On the other hand, travel rate exhibits minimal effect in the number of newly-infected individuals for both models. So long as NPIs are observed properly, mobility can be an option during pandemics.

Mathematical models in this study were tested for different combinations of NPIs to determine efficacy of each. Application of the social distancing measure on our simulations proved to be an effective measure to decrease the number of newly infected individuals. The best scenario would still be implementation of both social distancing and lockdown within cities. Relatively, absence of both NPIs would generate the most number of new infections, and is highly discouraged.

Both the ABM and compartmental (SEIR) model were favorable in simulating the transmission of infectious respiratory diseases in neighboring cities. Their simulations both exhibited akin behaviors when varying the parameters since both models are designed after each other. However, discrepancies in estimates occurred on the NPI comparisons for ABM and SEIR due to the heterogeneous and homogeneous mixing of individuals, respectively. Due to minimal heterogeneity and stochasticity effects, SEIR generated higher estimates of infected population compared to the ABM. ABM took a longer simulation time to reach similar outbreak values as SEIR since ABM considers both spatial and temporal aspect of the outbreak, and each individuals (agents) has their own stochastic characteristic and interaction.

Overall, our study demonstrated that between two cities, practice of non-pharmaceutical interventions, such as social distancing and lockdown, are necessary to reduce the risk of infection. With the practice of preventive measures, isolation of infected individuals and protection level of 75-100% effectiveness are ideal set-up to inhibit transmission of respiratory infectious diseases, such as SARS-CoV-2, especially when community lockdown rules are to be relaxed. Policy makers can use the results of both models in designing infectious disease-related policies to protect individuals while continuing population movement.

## 5. Limitations

The model considers selected dynamics of individuals only when inside the two cities. Transmission of disease is done through a distance function, and all infected individuals are assumed to recover after 14 days of infection. Death is not considered in the analysis of both model simulation results as the study focuses on the transmission of the disease only. Results of the study show the minimal effect of mobility during disease outbreaks but only through strict implementation and observance of the non-pharmaceutical interventions.

## 6. Supplementary File

The NetLogo file can be found online at: https://github.com/alvinizer/COVID19NLogoSimulations and a sample simulation can be viewed at: https://youtu.be/7q8twbpqQyU

## Data Availability

Information about data is included in the manuscript. The authors may be contacted for more details.

## Acknowledgment

JFR is supported by the Associate Scheme of the Abdus Salam International Centre for Theoretical Physics, Italy. Authors are members of the University of the Philippines COVID-19 Pandemic Response Team.

The authors declare no competing interests.

